# Role of the atmospheric pollution in the Covid-19 outbreak risk in Italy

**DOI:** 10.1101/2020.04.23.20076455

**Authors:** Daniele Fattorini, Francesco Regoli

## Abstract

After the initial outbreak in China, the diffusion in Italy of SARS-CoV-2 is exhibiting a clear regional trend with Northern areas being the most affected in terms of both frequency and severity of cases. Among multiple factors possibly involved in such geographical differences, a role has been hypothesized for atmospheric pollution. We provide additional evidence on the possible influence of air quality, particularly in terms of chronicity of exposure on the spread viral infection in Italian regions. Actual data on COVID-19 outbreak in Italian provinces and corresponding long-term air quality evaluations, were obtained from Italian and European agencies, elaborated and tested for possible interactions. Our elaborations reveal that, beside concentrations, the chronicity of exposure may influence the anomalous variability of SARS-CoV-2 in Italy. Data on distribution of atmospheric pollutants (NO_2_, O_3_, PM_2.5_ and PM_10_) in Italian regions during the last 4 years, days exceeding regulatory limits, and years of the last decade (2010-2019) in which the limits have been exceeded for at least 35 days, confirmed that Northern Italy has been constantly exposed to chronic air pollution. Long-term air-quality data significantly correlated with cases of Covid-19 in up to 71 Italian provinces (updated 27 April 2020) providing further evidence that chronic exposure to atmospheric contamination may represent a favourable context for the spread of the virus. Pro-inflammatory responses and high incidence of respiratory and cardiac affections are well known, while the capability of this coronavirus to bind particulate matters remains to be established. Atmospheric and environmental pollution should be considered as part of an integrated approach for sustainable development, human health protection and prevention of epidemic spreads but in a long-term and chronic perspective, since adoption of mitigation actions during a viral outbreak could be of limited utility.

**Capsule:** Chronic exposure to air pollutants might have a role in the spread of COVID-19 in Italian regions. Diffusion of Covid-19 in 71 Italian provinces correlated with long-term air-quality data.

## Main text

In December 2019, several pneumonia cases were suddenly observed in the metropolitan city of Wuhan (China), as the result of infection to a novel coronavirus (Li et al., 2020; Wu et al., 2020; Xu et al., 2020). This virus was termed SARS-CoV-2 for its similarity with that responsible of the global epidemic Severe Acute Respiratory Syndrome (SARS) occurred between 2002 and 2003 (Xu et al., 2020). Patients affected by SARS-CoV-2 infection often experienced serious complications, including organ failure, septic shock, pulmonary oedema, severe pneumonia and acute respiratory stress syndrome which in several cases were fatal (Chen et al., 2020; Sohrabi et al., 2020). The most severe symptoms, requiring intensive care recovery, were generally observed in older individuals with previous comorbidities, such as cardiovascular, endocrine, digestive and respiratory diseases (Sohrabi et al., 2020; Wang D. et al., 2020). The World Health Organization (WHO) has defined this new syndrome with the acronym COVID-19 for Corona Virus Disease 2019 (Sohrabi et al., 2020; WHO, 2020a).

The drastic containment measures adopted by Chinese government did not prevent the diffusion of SARS-CoV-2, which in a few weeks has spread globally. Italy was the first country in Europe to be affected by the epidemic COVID-19, with an outbreak even larger than that originally observed in China (Fanelli and Piazza, 2020; Remuzzi and Remuzzi, 2020). Other European countries and United States rapidly registered an exponential growth of clinical cases, leading to restrictions and a global lockdown with evident social and economic repercussions (Cohen and Kupferschmidt, 2020; ECDC, 2020). The WHO has recently declared the pandemic state of COVID-19 with over 2.8 million of cases reported and over 201.000 victims worldwide (Cucinotta and Vanelli, 2020; WHO, 2020b; ECDC, 2020, accessed on 27 April 2020).

The ongoing epidemic trend in Italy immediately showed strong regional differences in the spread of infections, with most cases concentrated in the north of the country (Remuzzi and Remuzzi, 2020). The distribution of positive cases reported from February 24 ^th^ to April 27 ^th^ is summarized in Figure 1A: some areas of Lombardy and Piedmont clearly exceeded 10.000 cases, e.g. 18.371 at Milan, 12.564 at Brescia, 11.113 at Bergamo, 12199 at Turin (data re-elaborated from the official daily reports of the Department of Civil Protection, ICPD, 2020, accessed on 27 April 2020). Also, the relative percentage distribution of the positive test rate (Figure 1B) exhibit higher values in Northern Italy despite a certain uncertainty of data due to the different numbers and frequency of oropharyngeal swabs performed in various regions to test coronavirus positivity; mortality rate ranged from 18% in the most affected, northern regions to less than 5% in the others (Figure 1C). Overall these trends are confirmed from the rates of reported COVID-19 cases and of fatal events, expressed as percentage values normalized to the number of inhabitants for regional populations (Figures 1D and 1E), further corroborating significantly greater effects in Northern Italy, both in terms of number of infections and the severity of cases (mortality).

**Figure 1.**
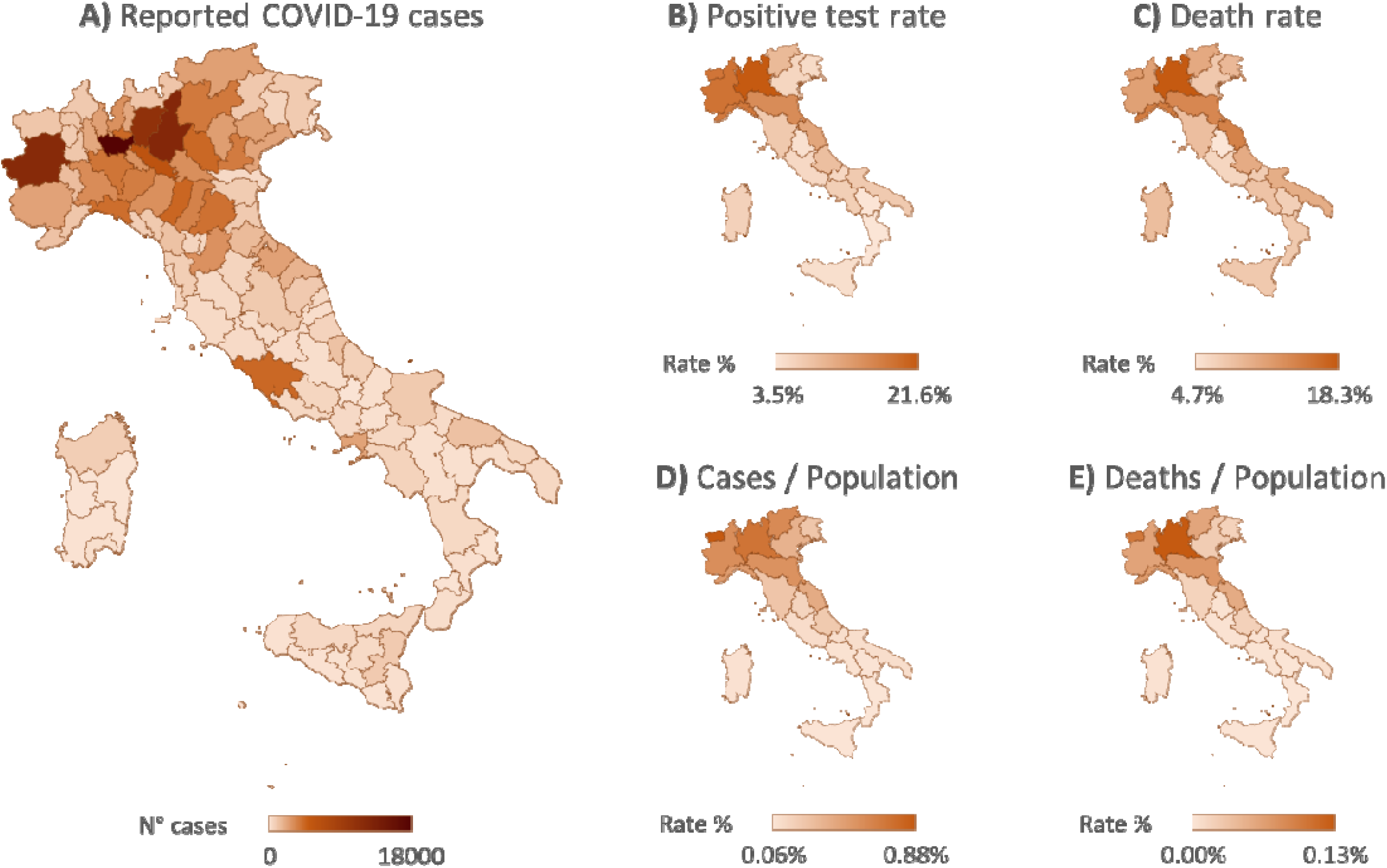
Regional distribution of COVID-19 outbreak in Italy (from 24 February to 27 April 2020. A) abundance of COVID-19 cases (absolute number); B) percentage of positive subjects referred to the number of performed tests (oropharyngeal swabs); C) mortality rate on the number of positive cases; D) percentage of COVID-19 cases normalized to the number of inhabitants; E) percentage of deaths normalized to the number of inhabitants. Data obtained and re-elaborated from the official daily reports of Italian Civil Protection Department (ICPD, 2020).

To explain such geographical trend, it was initially assumed that restrictions decided by government authorities after the first outbreak in Lombardy, had contained the effects of the infection, preventing its rapid spread to the rest of the country. Some authors, however, from the clinical course of a large cohort of patients, have concluded that the epidemic coronavirus had been circulating in Italy for several weeks before the first recognized outbreak and the relative adopted containment measures (Cerada et al., 2020). In this respect, the differentiated occurrence of infection cannot be fully explained by the social confinement actions. Our elaboration we did not include the analysis of other possible determinants of incidence and mortality, such as age structure, capacity of the healthcare system, duration of the confinement since some of these data are currently not immediately available. Future studies will be necessary to fill such gap of knowledge addressing at which extent all these factors may have mutually contributed to the diffusion of COVID-19 in Italy. However, the current profile of the viral outbreak in Northern Italy suggests that other factors could be involved in the diffusion of infection and mortality rates.

Since the presence of comorbidities appeared determinant for the aetiology and severity of the COVID-19 symptoms (Chen et al., 2020; Wang T. et al., 2020; Wu et al., 2020), the role of atmospheric pollution in contributing to the high levels of SARS-CoV-2 lethality in Northern Italy has been hypothesized (Conticini et al., 2020). Association between short-term exposure to air pollution and COVID-19 infection has been described also for the recent outbreak in China (Zhu et al., 2020). The adverse effects of air pollutants on human health are widely recognized in scientific literature, depending on various susceptibility factors such as age, nutritional status and predisposing conditions (Kampa and Castanas, 2008). Chronic exposure to the atmospheric pollution contributes to increased hospitalizations and mortality, primarily affecting cardiovascular and respiratory systems, causing various diseases and pathologies including cancer (Brunekreef and Holgate, 2002; Kampa and Castanas, 2008). Among air pollutants, the current focus is mainly given on nitrogen dioxide (NO_2_), particulate matter (PM_2.5_ and PM_10_) and ozone (O_3_), frequently occurring at elevated concentrations in large areas of the planet.

The percentage of European population exposed to levels higher than the regulatory limits is about 7-8% for NO_2_, 6-8% for PM_2.5_, 13-19% for PM_10_ and 12-29% for O_3_ (EEA, 2019). Premature deaths due to acute respiratory diseases from such pollutants are estimated to be over to two million per year worldwide and 45.000 for Italy (Brunekreef and Holgate, 2002; Huang et al., 2016; EAA, 2019; Watts et al., 2019).

Here, we are providing additional evidence on the possible influence of air quality on the spread of SARS-CoV-2 in Italian regions. Since the effects of air pollutants on human health not only depend on their concentrations but also, if not especially, on chronicity of exposure, we have elaborated the last four years (from 2016 to 2019, EEA, 2020) of regional distribution of NO_2_, PM_2.5_ and PM_10_ is presented in Figure 2. The highest atmospheric concentrations were clearly distributed in the Northern areas (Piedmont, Lombardy, Veneto and Emilia-Romagna), in addition to urbanized cities, such as Rome and Naples.

**Figure 2.**
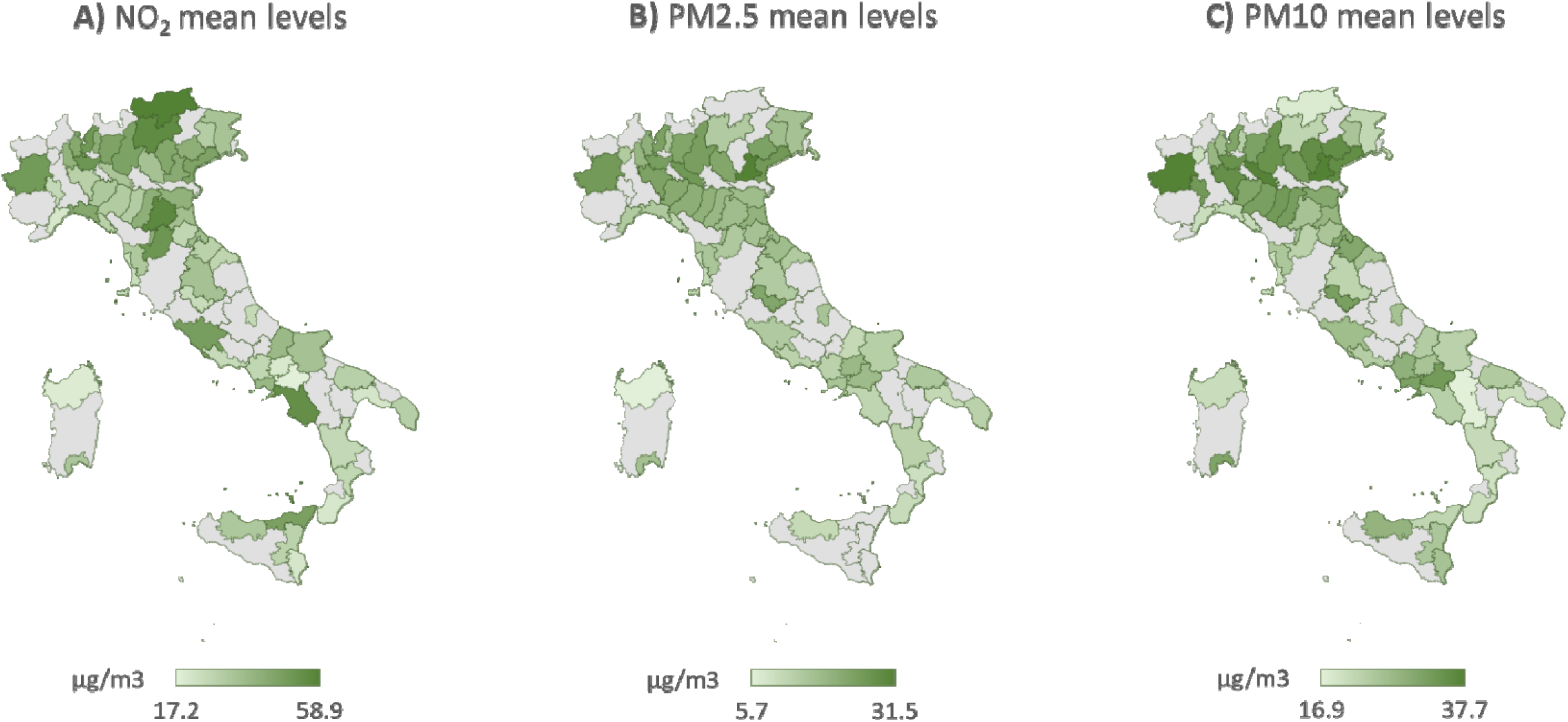
Regional data on air quality levels: A) nitrogen dioxide (NO2); B) particulate matter of 2.5 µm or less (PM2.5); C) particulate matter of 2.5-10 µm^3^ (PM10). Data are referred to the means of values of last four years (2016-2019), expressed as µg/m^3^ (obtained and elaborated from the European Environmental Agency, accessed on 6 April, EEA, 2020).

The long-term condition of population exposure is also revealed by the number of days per year in which the regulatory limits of O_3_ and PM_10_ are exceeded (Figure 3A-B): the critical situation of Northern Italy is reflected by values of up to 80 days of exceedance per year (average of the last three years, EAA, 2019). Worthy to remind, ozone is one of the main precursors for the formation of NO_2_, and chronic exposure to this contaminant for almost a quarter of a year is undoubtedly of primary importance. The chronic air pollution in Northern Italy is further represented by the number of years during the last decade (2010-2019) in which the limit value for PM_10_ (50 µg/m^3^ per day) has been exceeded for at least 35 days (Figure 3C). Once again, these data confirm that the whole Northern area below the Alpine arc has been constantly affected by significantly higher levels of these contaminants.

**Figure 3.**
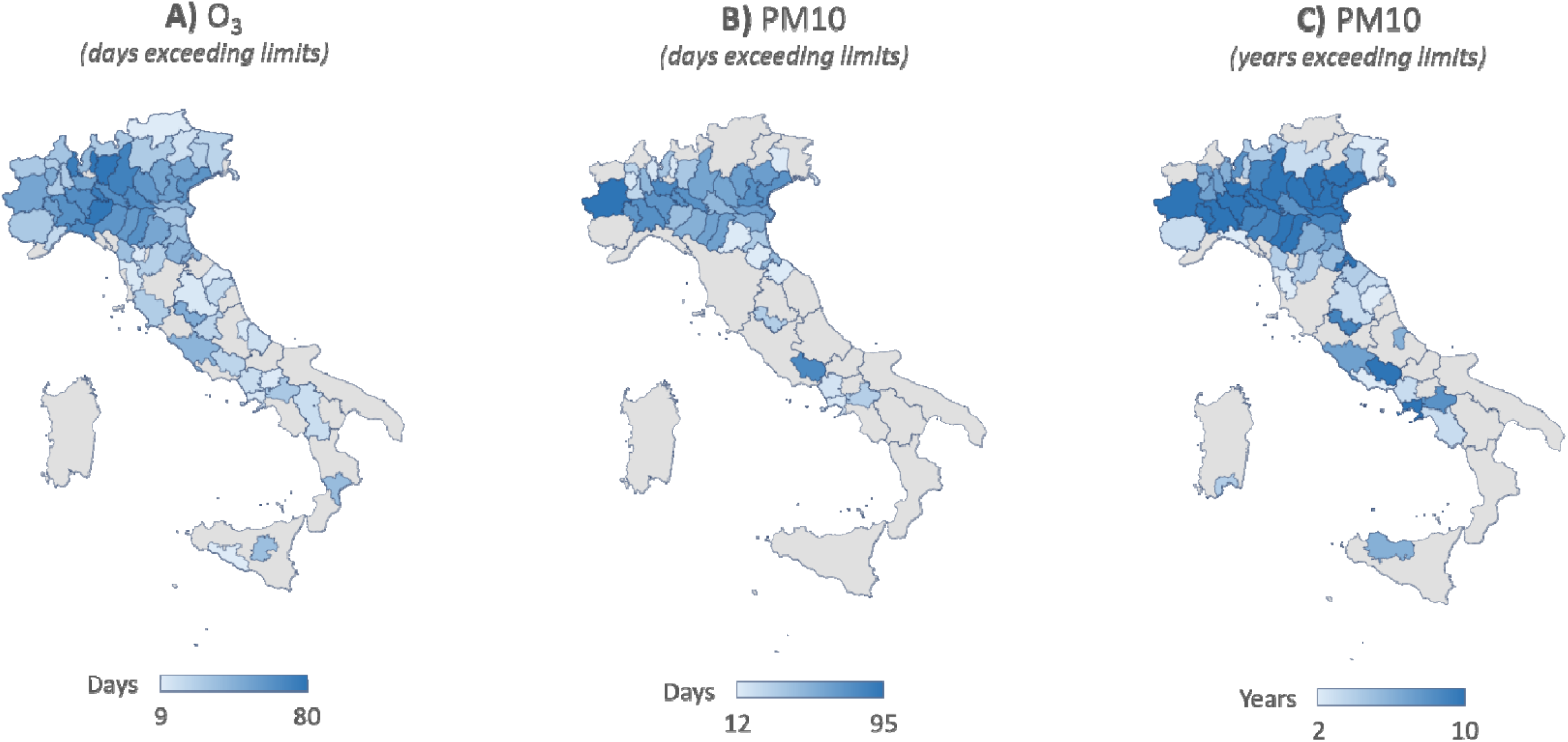
Number of days per year exceeding the regulatory limits relating to A) ozone (O3) and to B) particulate matter (PM10), as average means of the last 3 years (2017-2019); C) number of years in which the PM10 limit was exceeded for at least 35 days per year, from 2010 to 2019. Data are obtained and elaborated from annual reports (Legambiente, 2018; 2019; 2020) and referred to the official statistics of the European Environmental Agency (EEA, 2019).

The hypothesis that atmospheric pollution may influence the SARS-CoV-2 outbreak in Italy was also tested from the relationships between the confirmed cases of Covid-19 in up to 71 Italian provinces (updated 27 April 2020) with the corresponding air quality data. The latter were expressed as average concentrations in the last 4 years of NO_2_, PM_2.5_ and PM_10_ (Figure 4A-C) and the number of days exceeding the regulatory limits (averages of the last 3 years) for O_3_ and PM_10_ (Figure 4D-E). The always significant correlations provided further evidence on the role that chronic exposure to atmospheric contamination may have as a favourable context for the spread and virulence of the SARS-CoV-2 within a population subjected to a higher incidence of respiratory and cardiac affections.

**Figure 4.**
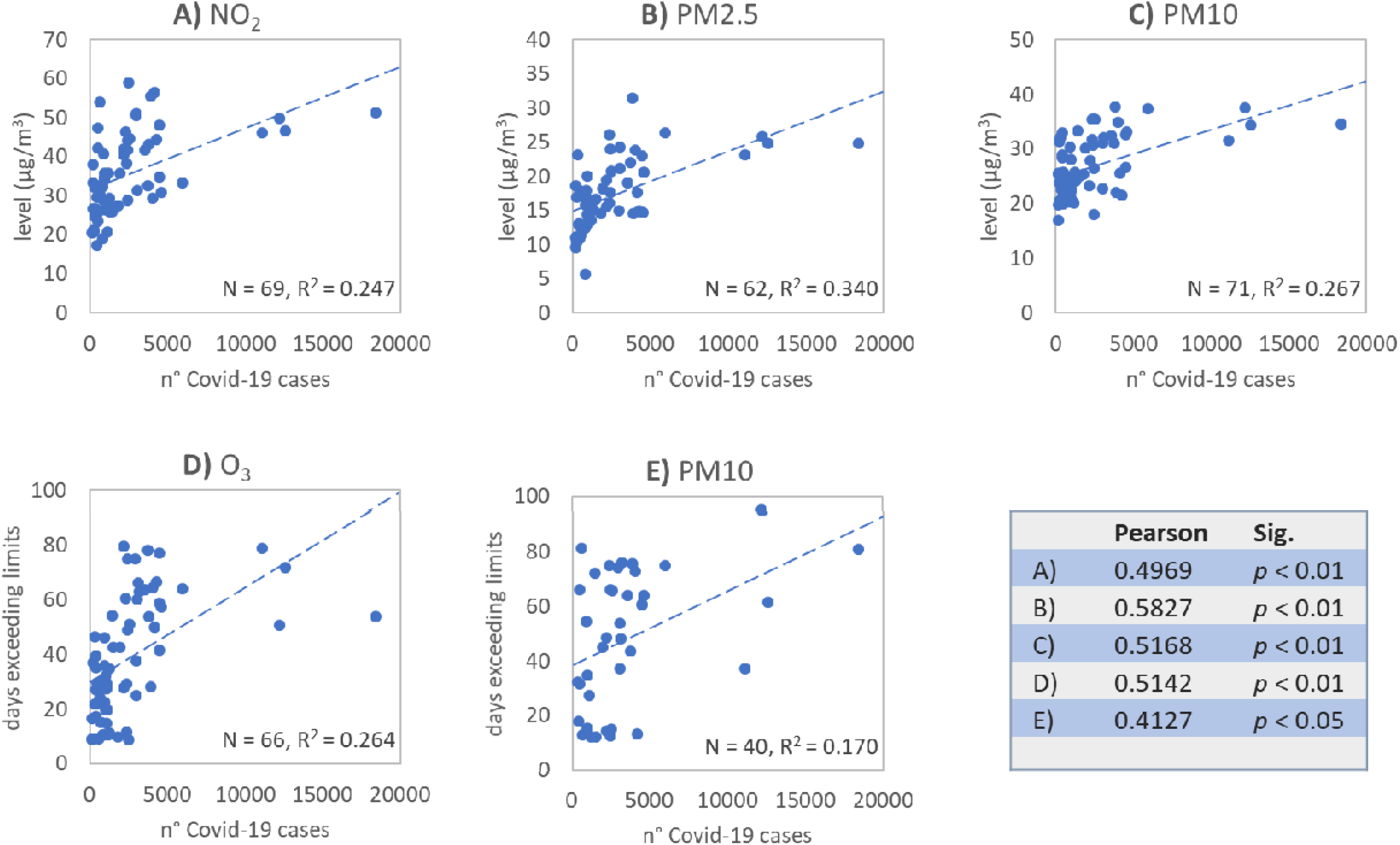
Statistical correlation between the regional distribution of COVID-19 cases and the air quality parameters in Italy: incidence of COVID-19 cases vs levels of A) NO2, B) PM2.5 and C) PM10 (four years means); incidence of COVID-19 cases vs number of days exceeding regulatory limits of D) O3 and E) PM10 (three years means). Data obtained and elaborated from EEA (2019; 2020), ICPD (2020), and Legambiente (2018; 2019; 2020).

It is well known that exposure to atmospheric contaminants modulate the host’s inflammatory response leading to an overexpression of inflammatory cytokines and chemokines (Gouda et al., 2018). Clear effects of Milan winter PM_2.5_ were observed on elevated production of interleukin IL-6 and IL-8 in human bronchial cells (Longhin et al., 2018), and also NO_2_ was shown to correlate with IL-6 levels on inflammatory status (Perret eta al., 2017). The impairment of respiratory system and chronic disease by air pollution can thus facilitate viral infection in lower tracts (Shinya et al., 2006; van Riel et al., 2006).

In addition, various studies have reported a direct relationship between the spread and contagion capacity of some viruses with the atmospheric levels and mobility of air pollutants (Ciencewicki and Jaspers, 2007; Sedlmaier et al., 2009). The avian influenza virus (H5N1) could be transported across long distances by fine dust during Asian storms (Chen et al., 2010), and atmospheric levels of PM_2.5_, PM_10_, carbon monoxide, NO_2_ and sulphur dioxide were shown to influence the diffusion of the human respiratory syncytial virus in children (Ye et al., 2016), and the daily spread of the measles virus in China (Chen et al., 2017; Peng et al., 2020).

These evidences and presented elaborations would confirm that the anomalous variability of the diffusion and virulence of SARS-CoV-2 in Italy could partly depend on the levels of atmospheric contamination. Although the capability of this coronavirus to bind particulate matters remains to be established, chronic exposure to atmospheric contamination and related diseases may represent a risk factor in determining the severity of COVID-19 syndrome and the high incidence of fatal events (Chen et al., 2020; Conticini et al., 2020; Dutheil et al., 2020; Wang D. et al., 2020; Wu et al., 2020).

In conclusion, the actual pandemic event is demonstrating that infectious diseases represent one of the key challenges for human society. Since periodic emergence of viral agents show an increasing correlation with socio-economic, environmental and ecological factors (Morens et al., 2004; Jones et al., 2008), also air quality should be considered as part of an integrated approach toward sustainable development, human health protection and prevention of epidemic spreads. However, the role of atmospheric pollution should be considered in a long-term, chronic perspective, and adoption of mitigation actions only during a viral outbreak could be of limited utility.

## Data Availability

With the present I certify the availability of all the data used and elaborated in the paper, at the links given below.

https://github.com/pcm-dpc/COVID-19

https://www.eea.europa.eu/publications/air-quality-in-europe-2019

https://www.eea.europa.eu/themes/air/air-quality-and-covid19/monitoring-covid-19-impacts-on

https://www.legambiente.it/malaria-di-citta/

## Acknowledgement

No funding was received for this study.

## Note

*A complete description of the origin of the used data and the methods of graphic and statistical processing is included in the Supplementary Materials*.

---

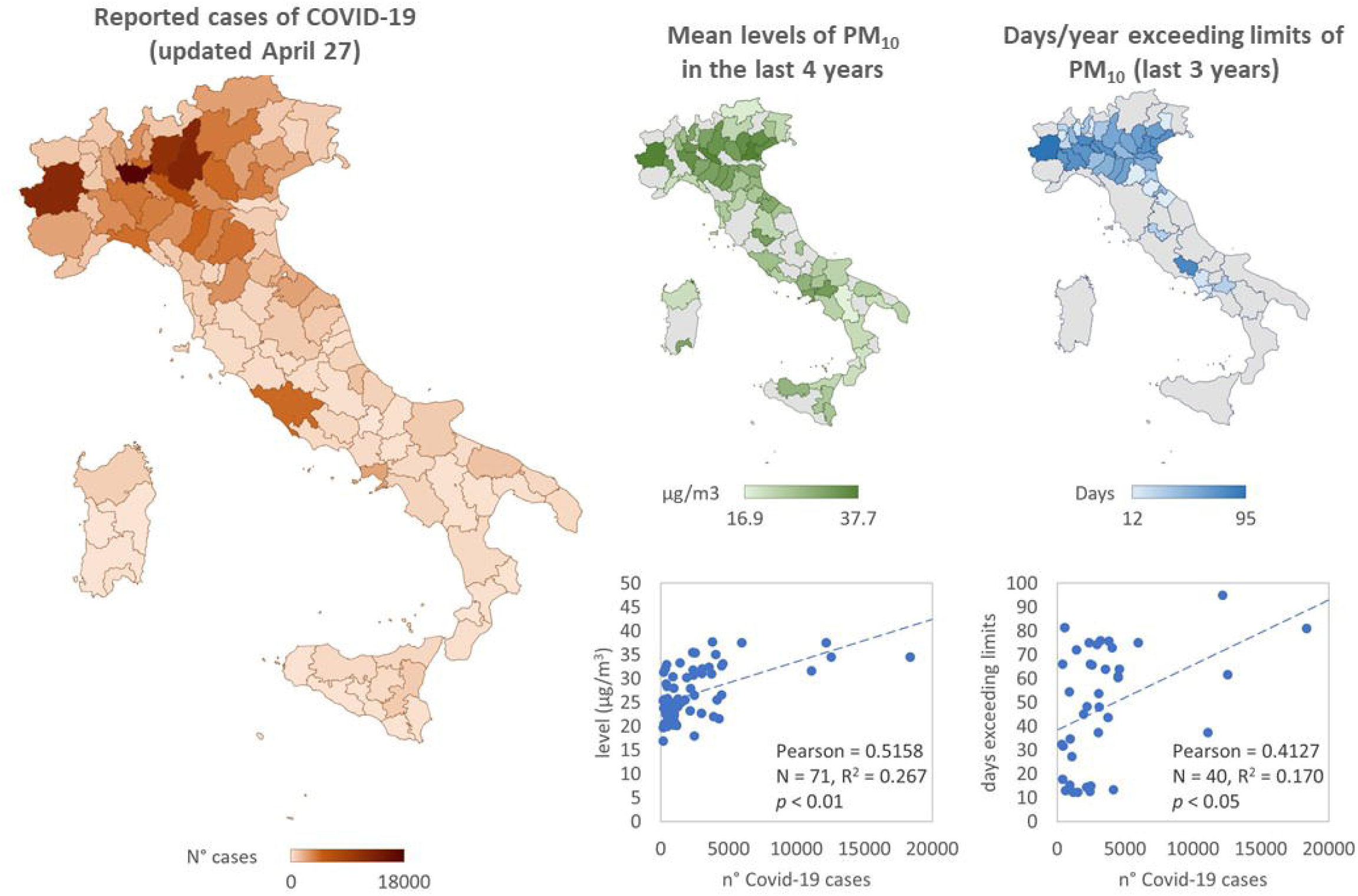

## Notes

### Competing Interest Statement

The authors have declared no competing interest.

### Funding Statement

No external funding was received to elaborate presented data

